# Estimating rates of SARS-CoV-2 lineage spread from graph theory analysis and data mining of genetic sequence data streams

**DOI:** 10.1101/2025.02.07.25321892

**Authors:** Emma E. Goldberg, Michael D. Kupperman, Ruian Ke

**Affiliations:** Theoretical Biology and Biophysics, Los Alamos National Laboratory, Los Alamos NM, USA; Deparment of Applied Mathematics, University of Washington, Seattle WA, USA

## Abstract

The modern enormous scale of viral genomic data, as exemplified by SARS-CoV-2, presents unique opportunities for epidemic inference and real-time monitoring. Existing methods, however, cannot efficiently process the extremely large stream of sequence data and simultaneously identify small subsets of sequences that may represent rapid population growth of newly emerged lineages. To address this gap, we developed a new approach that combines techniques in traditional graph theory and modern data mining. We represent small subsets of sequences as graph Laplacians and identify from them features of rapid population growth. From early sequence data collected in the US and the UK between 2021 and mid-2022, we identified two features—genetic diversity and matrix connectivity—that allow us to reliably estimate growth rates of newly emerged lineages. To test our model, we used data collected from mid-2022 and end-2024 and accurately predicted the growth rates of lineages that appeared during this period. Furthermore, for data collected in 2023 when the sequencing efforts were relatively high (thousands of sequences per day) in the US and the UK, our model correctly identified the most rapidly expanding lineages when they were still at low frequencies (between 1-6%). Overall, our work provides a scalable and adaptable tool to estimate the growth rate of newly emerged SARS-CoV-2 lineages. More broadly, the interpretable logic of our method suggests potential for rapid outbreak identification for other rapidly evolving pathogens.

**Significance Statement:** The COVID-19 pandemic has underscored the importance of rapidly identifying emerging SARS-CoV-2 lineages that may cause large outbreaks. Methods are needed that can handle a large amount of incoming genomic data and at the same time identify small groups of cases that may represent rapid growth of newly emerged lineages. To address this need, we developed an approach utilizing graph theory and data mining techniques, which we show reliably estimates the growth rates of newly emerged SARS-CoV-2 lineages. Our approach can promptly identify rapidly growing lineages, often weeks or months before they became dominant. This demonstrates its potential as a scalable tool for transmission dynamics inference, real-time outbreak monitoring, and pandemic preparedness.

## Introduction

Since the start of the COVID-19 pandemic, variants of SARS-CoV-2 with higher transmissibility and/or ability to escape immunity have been continuously generated, and some of them, particularly the pre-Alpha, Alpha, Delta, and many Omicron variants, spread globally and caused major outbreaks [1, 2, 3, 4]. Such continuous viral evolution means that SARS-CoV-2 variants must be closely monitored, such that the rapidly expanding variants identified as early as possible. This would in turn facilitate further characterizations to understand public health consequences and formulate effective vaccines [5].

Viral genetic sequencing is frequently used in outbreak investigations [6], but computational tools utilizing these data for real-time monitoring have lagged behind data availability [5]. Rapidly expanding SARS-CoV-2 sequence availability poses unique opportunities and challenges for transmission dynamics inference and real-time monitoring. Thousands or tens of thousands of sequences per day means that an effective inference approach should be able to rapidly take advantage of a large amount of sequence data. Simultaneously, however, the need for reliable estimation of the growth rate and thus rapid identification of newly-emerged and quickly-expanding lineages from background requires an approach that can make predictions about a lineage using only a small amount of data associated with it.

Traditional approaches often fail to address these challenges jointly. For example, the rise in counts or frequency of a new variant can be used to infer its growth rate, and hence the potential of its outbreak [7, 8, 9]. However, there are inherent noise and biases in the time series due to under-sampling or reporting delay [9]. This means often a long time series is required for regression-based approaches, even up to when the variant attains a substantial frequency and hence the outbreak is beyond control [7]. We showed that a machine learning approach based on transformers can learn and mitigate the impacts of biases and noises in the time series [9], but variant-based approaches in general rely on correct and timely assignment of new lineages [10]. Phylodynamic models can infer epidemiological quantities such as the rate of spread and thus the basic reproductive number, often in a Bayesian inference framework that jointly infers phylogenetic trees and population histories [11, 12]. This approach works best with moderate amounts of data, however: with very few sequences early in an outbreak there is little power to fit parameter-rich models, and with very many sequences the computational cost is too high.

Most recently, methods using tree-free approaches based on statistical or machine learning techniques have been developed to handle large amounts of viral sequences [13, 14, 15]. For example, a model of epidemiological dynamics has been used with a simple summary statistic of sequence data (the number of segregating sites in a sample) to estimate the transmission dynamics of new SARS-CoV-2 variants [13]. Another work used the distribution of the size of identification sequence clusters to estimate the effective reproductive number and the dispersion parameter [15]. These studies demonstrated the potential utility of summary statistics of sequence data in transmission dynamics inference. They are computationally more tractable than phylogenetic analyses; however, reliable inference still require a substantial amount of data and may depend on other parameters assumed in the transmission and reporting processes. We recently developed and trained a deep neural network model to identify HIV outbreak clusters from sequence data [14]. This approach treats pairwise distance matrices of subsets of sequences as images and trains a convolutional neural network model to identify signals of outbreak in the images. Since the model is trained on data generated from an HIV transmission simulator using parameters estimated from extensive prior datasets, this approach is also likely to rely on accurate specification of transmission model parameters.

Here, we present a tree-free and computationally efficient approach to analyze the SARS-CoV-2 genomic sequences using graph-theory and data mining techniques. This approach leverages the large amount of SARS-CoV-2 genomic data by deriving features from lower-dimension representation of the sequences and estimating the rate of lineage expansion directly from the data without additional assumptions on transmission dynamics. Below, we present an overview of our approach first, and then describe the analyses of SARS-CoV-2 genomic data and the identification of two graph features that allow for reliable prediction of the rate of lineage expansion. At the end, we perform retrospective analyses to demonstrate our approach can readily identify the most rapidly growing newly-emerged lineages from an evolving genomic database.

## Results

### Overview of our approach

Our premise is that the expansion rate of a newly emerged SARS-CoV-2 lineage can be estimated by analyzing summary statistics and graph-theory features derived from sets of closely related viral genomic sequences collected during the outbreak. This approach uses and analyzes pairwise distance matrices that are formed from sets of closely-related sequences in the database. As we shown in our previous work [14]), this allows for efficient processing the large amount of SARS-CoV-2 sequence data available in the database. We then develop a novel method to derive features of these matrices by first converting the pairwise distance matrices to graph Laplacian, such that each node in the graph represents a sequence and the weight of the edge in the graph is given by the distance between two corresponding sequences. We then analyze the eigen-spectra of the Laplacians. Our approach is founded on the idea that because the eigenspectrum of a graph Laplacian represents aspects of its connectivity between nodes in the graph [16], it thus also reflects the topological structure of the sequences in the space represented by their genetic distances. Since it has been well-established that the topological structure (e.g., represented by phylogenetic trees) is heavily influenced by past population dynamics [17, 18], we reason that the eigenspectrum should contain features of the viral transmission history in the near past. We utilize past SARS-CoV-2 lineage dynamics and sequence data to derive such features and show that these features provide strong predictive power for promptly estimating the growth rate of a newly emerged lineage using only a small amount of sequences.

Our approach proceeds in three steps. First, we formed pairwise distance matrices of small sets of closely-related sequences (‘submatrices’ of 50 sequences each) taken from fourteen Pango lineages that became highly successful in the US and the UK before mid-2022, including time periods of both their growth and decline (Fig. 1A and B). We converted these matrices to graph Laplacians, and performed principal components analysis (PCA) on their eigenspectra (Fig. 1C), finding statistical features that differentiate lineage growth from decline (Fig. 1D and E). Second, we applied the PCA projections to matrices from a much larger collection of Pango lineages in the UK and the US (between January 2020 and October 2022) to identify the graph features which were then used in a linear regression model to predict the rates of population change for each variant. This resulting regression model is easily interpretable as a combination of genetic diversity and connectivity within a small sample of closely-related viral sequences. Third, to test the predictive power of this regression model, we applied it to data collected after October 2022 (and therefore not used in training the model) to demonstrate that it can estimate epidemic growth rates and thus identify rapidly-growing lineages from early genetic data for each lineage.

**Figure 1:**
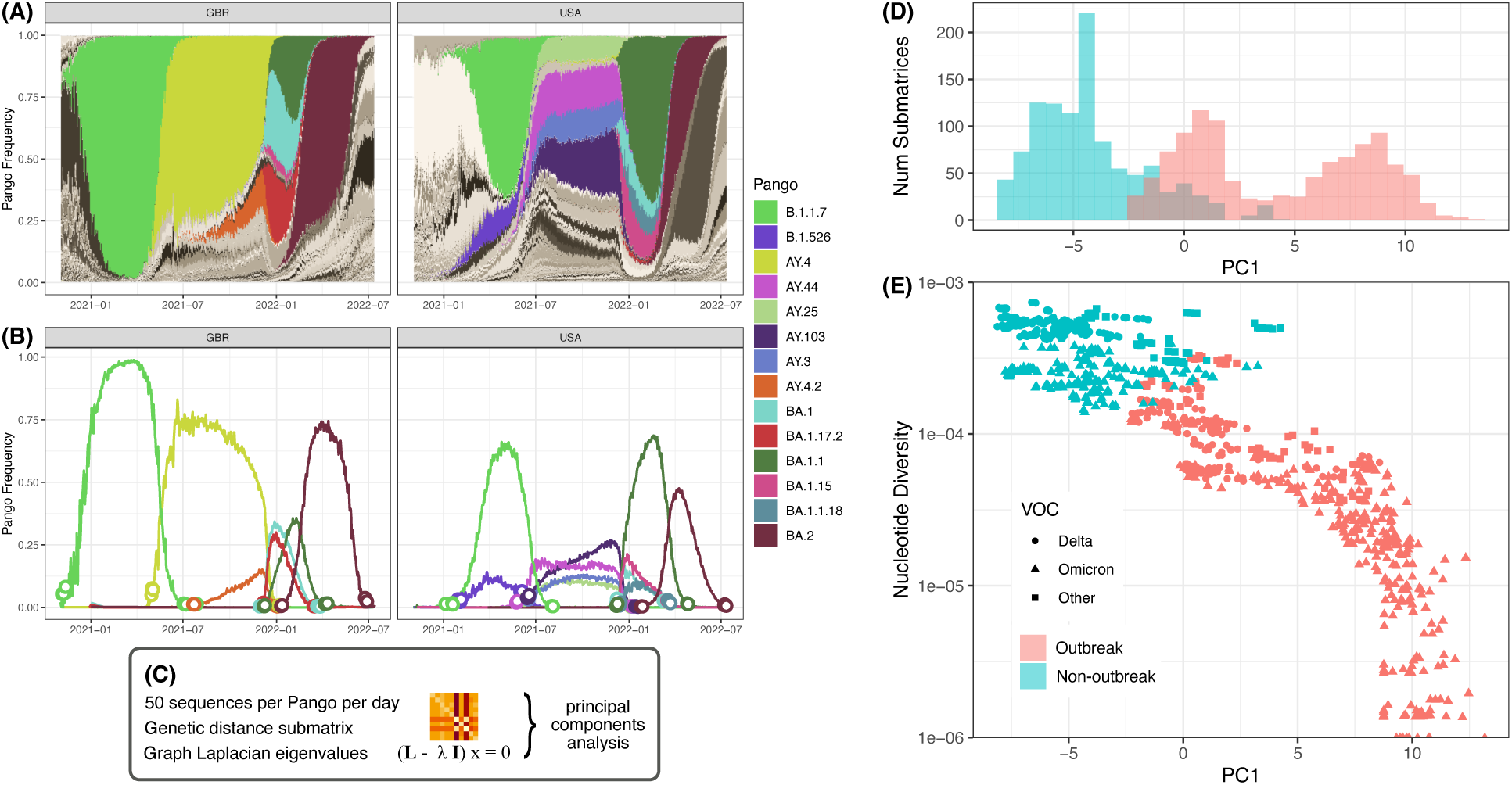
Graph Laplacian PCA to distinguish outbreak from non-outbreak. From all the Pango variants present in GBR and USA during the time period we used for training (A), we identified 14 that had sufficient data (for submatrix size 50) during clear expanding/outbreak and decline/non-outbreak phases, and reached a frequency of at least 10% (B). For each, we formed pairwise distance submatrices and their corresponding normalized graph Laplacian matrices during days of the early rise and late decline of the variant, computed their eigenvalues, and performed a PCA on all of the resulting values (C). We found that the first principal component (PC1) substantially distinguished the outbreak from non-outbreak data (D), and that additionally including the submatrix nucleotide diversity further distinguished the data and even the broader variant types (E).

Below we describe results from each step of the analysis. Further details are provided in Methods.

### SARS-CoV-2 genomic data collection and construction of pairwise distance matrices

We used SARS-CoV-2 metadata and genomic sequence data downloaded from GISAID [19] on 2024-10-07 and cleaned and aligned by the Nextclade pipeline [20]. For each combination of Pango lineage, country, and day with sufficient data, we formed 20 submatrices, each composed of the 50 sequences most closely related to one randomly-sampled sequence. These submatrices formed the basis of our subsequent analyses. The choice of submatrix size is based on our previous work predicting and identifying outbreaks using matrices of pairwise distances of HIV sequences [14]. Later in this work, we tested robustness of our conclusions against the choice of this number.

### Graph Laplacian PCA distinguishes outbreak from non-outbreak

To identify statistical features related to population growth rate, we first focused on the submatrices from the initial expansion (‘outbreak’ phase) and final decline (‘non-outbreak’ phase) for the subset of Pango lineages that reached high frequency in the US and the UK before July 2022 (Fig. 1B). We converted each submatrix into its corresponding graph Laplacian, with each sequence as a node in the graph and the pairwise genetic distance between two sequences representing the strength or weight of their connection. We then calculated the eigenspectra of the graph Laplacians, and performed PCA on the collection of eigenspectra from all the outbreak and non-outbreak Laplacian matrices to uncover the main features separating the two phases.

We found that the first principal component of the PCA correlates almost perfectly with the nucleotide diversity of the pairwise distance submatrices (Pearson’s *r* = -0.999; Fig. S1A). This is expected because of the data we used for training: the collection of sequences from the very early emergence of a new variant will contain little genetic diversity (relative to one another), while sequences from much later will contain much diversity. Consequently, the first principal component separates the outbreak from non-outbreak submatrices extremely well (explaining 99.5% of the variance; Fig. S1B, Fig. S2A). We therefore retained nucleotide diversity as one potential predictor of outbreak growth rate. However this measure, although simple to derive, does not capture more subtle explanatory aspects such as the connectivity of the sequences, and it may not be generally reliable because it can be affected by surveillance properties such as the level of sampling effort and the time when a lineage is detected.

Because the unnormalized Laplacian eigenspectrum is dominated by the magnitude of genetic diversity, we next examined the normalized Laplacian, in which the genetic distances are adjusted to remove the impact of the magnitude of genetic diversity, while the relatedness of each sequence to other sequences is preserved. This is a means to seek additional aspects of outbreak structure in the graph Laplacians that could help to predict growth rate. We found that a large fraction (61%) of the variation in the eigenspectrum of the normalized Laplacian is explained by the first principal component (Fig. S2B), and that this component distinguishes quite clearly the outbreak from non-outbreak submatrices (Fig. 1D). This is remarkable given that the overall level of genetic diversity no longer contributes, and that PCA, as an unsupervised algorithm, is agnostic with respect to whether a submatrix belongs to an outbreak versus non-outbreak scenario.

Finally, we found that the combination of PC1 from the normalized Laplacian (hereafter, ‘PC1’) and the submatrix nucleotide diversity even more clearly separate the outbreak and non-outbreak data, and even somewhat distinguish the main outbreaking lineages (Delta and Omicron from others; Fig. 1E). In general, matrices from the more rapidly expanding variants had lower genetic diversity and a larger PC1 value. For example, submatrices from the most rapidly expanding Omicron variant mostly clustered at the bottom-right corner of the map (in Fig. 1E). Intuitively, the more rapid the outbreak, the smaller the number of genetic mutations by the time sufficiently many samples are available. Additionally, more rapid outbreaks leave distinct patterns in a pairwise distance matrix [14] and thus different PC1 values.

### Graph features of pairwise distance submatrices explain the growth/decline rate

Based on the results above, we hypothesized that these features derived from the pairwise distance submatrices— the nucleotide diversity and PC1 from the normalized graph Laplacian—can be used to estimate the rate of epidemic growth of a newly emerged lineage. To test this hypothesis, we first estimated the rates of growth or decline for each lineage by fitting a piecewise linear regression (PWLR) model to the log-transformed variant frequency time series (Fig. 2A). We use the estimated rates to represent the ‘true’ rates of growth or decline below. Note that this estimation is only possible retrospectively, because it requires data collected both before and after submatrices were collected. Second, for each lineage (in each country) we calculated the mean of PC1 and the logarithm of the nucleotide diversity during the first 3-day window (representing the initial outbreak phase) and the last 3-day window (representing the non-outbreak decline phase) when 50 or more sequences per day were available.

**Figure 2:**
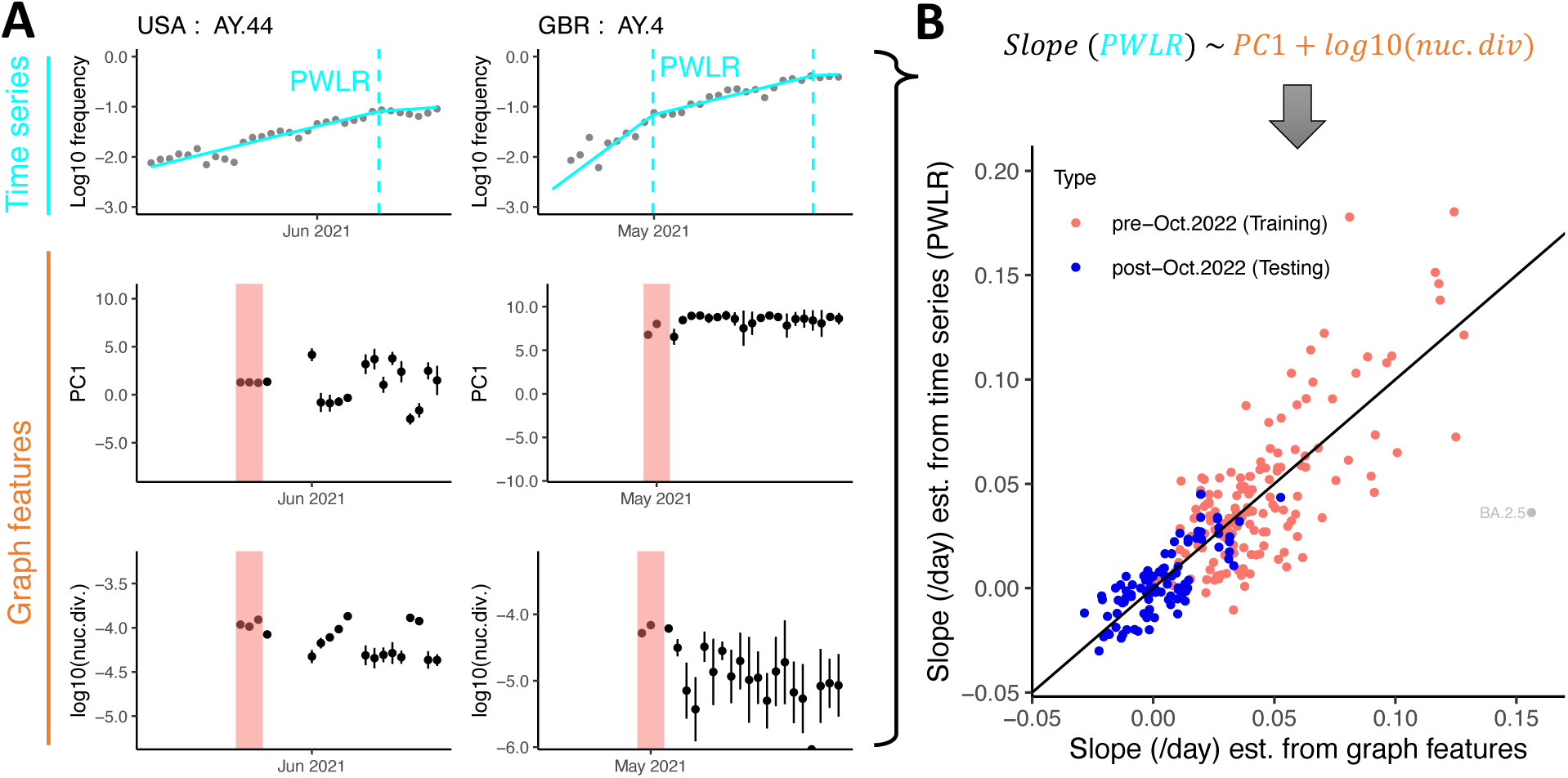
Linear regression model combining graph connectivity (PC1) and nucleotide diversity (nuc.div) to predict lineage growth rate (slope). (A) For each lineage in each country (Delta variants AY.44 in USA and AY.4 in the UK shown here as examples), we fit a piecewise linear regression (PWLR, cyan) to the log10 of the the frequency timeseries to obtain the ‘true’ rates of lineage growth (slopes). We then compute graph features from submatrices (Fig. 1CDE) when they become available for each lineage (pink bars). (B) The graph features were used as independent variables and the PWLR slopes as dependent variables in a linear regression model. The slopes predicted from this model (horizontal axis) correlated well the slopes from the PWLR (vertical axis). Pink dots represent the training data used to infer parameter values in the regression analysis, whereas blues dots represent the testing data used to test the regression predictions. BA.2.5 from the UK (gray dot) were excluded from the regression because of extremely low genetic diversity.

To evaluate how well the two features explain and predict the ‘true’ growth and decline rates, we separated them into pre-Oct. 2022 and post-Oct. 2022 groups, which were then used as training and testing datasets, respectively. For model training, we performed a multivariate regression first using data from both the outbreak and the non-outbreak phases (Fig. S4). Because there is a lack of spread and a large amount of noise in the decline rate during non-outbreak phases, we also performed the same regression using data from the outbreak phase only (Fig. 2B). In both cases, the multivariate regression model estimates agree very well with the ‘true’ rates for nearly all lineages (Fig. 2B and Fig. S4). An exception is BA.2.5 in UK, which has extremely low nucleotide diversity (< 3 ⇥ 10^-6^; Fig. S3), meaning most of the sequences from BA.2.5 are identical, and leading to an overestimate of its growth rate. We therefore excluded BA.2.5 from our analysis.

Then, the combination of log10 of nucleotide diversity and PC1 explain 70% and 56% of variation in the slope (*R*^2^ values of 0.70 and 0.56, see Fig. 2B and Fig. S4) for the model using data from both phases and the model using data from the outbreak phases only, respectively. Note that the two percentages are not directly comparable because the number of data points differ.

We then used the testing dataset to test how well the trained regression models make predictions. The resulting residual least squares errors are 0.042 and 0.011 for regressions using data from both phases and data from outbreak phases only, respectively. It is clear that the model using data from both phases tends to underestimate the rates in the testing sets, while the model using data from the outbreak phase only predicts the rates better (compare Fig. S4 to Fig. 2B). This is likely due to the higher level of noise in the rate of change from the non-outbreak phase data. Therefore, we use the regression model using data from the outbreak phase only in the analysis below to identify rapidly growing new variants.

### Correct identification of the rise of future dominating variants

To demonstrate the utility of our approach in informing real-time SARS-CoV-2 variant surveillance, we retrospectively evaluated how well our regression model identifies rapidly-growing and future-dominating lineages among all extant lineages. We selected several time periods between when a dominating lineage first appeared in the population and when it became dominant, and we derived the submatrix graph features for all lineages with at least 50 sequences over during these periods. We estimated the slope of lineage growth from the two graph features (Fig. 2B), and tested if our model identified the future-dominating lineages as the most rapidly growing ones when they emerged in the population.

We firsts applied our model to data during the periods when the training data were collected. As expected, our model correctly identified the lineages that eventually came to dominate (Alpha, Delta, and Omicron BA.1 variants) as the most rapidly expanding ones in the UK and the US (Figs. S7 and S8). For Alpha and Delta, we would be able to identify these lineages about one month before the lineage became dominant. Because of the rapid rise of BA.1 and thus the short period between its appearance and dominance, our model would identify BA.1 as the emerging dominant lineage 11 to 16 days in advance.

We then tested model performance using the same type of analysis on the testing data (collected after the time period used to train the model). The model correctly identified the dominating lineages (BQ.1, XBB.1.5, HV.1 and JN.1) in the US as soon as there were 50 sequences available in a day to form the submatrix (Fig. 3). The identification is made when each lineage was at a frequency between 1-6% and between 21 and 41 days before the lineages became the dominant lineage. For data from the UK, our model is also able to identify the dominant lineages (BQ.1.1, XBB.1.5 and JN.1) as soon as there are enough sequences to construct the submatrices (Fig. 4). However, because the sequencing effort in the UK was relatively low in 2023 and 2024 (tens to hundreds of sequences per day, Fig. S10), XBB.1.5 and JN.1 had already risen to a high frequency at the time of identification.

**Fig. 3:**
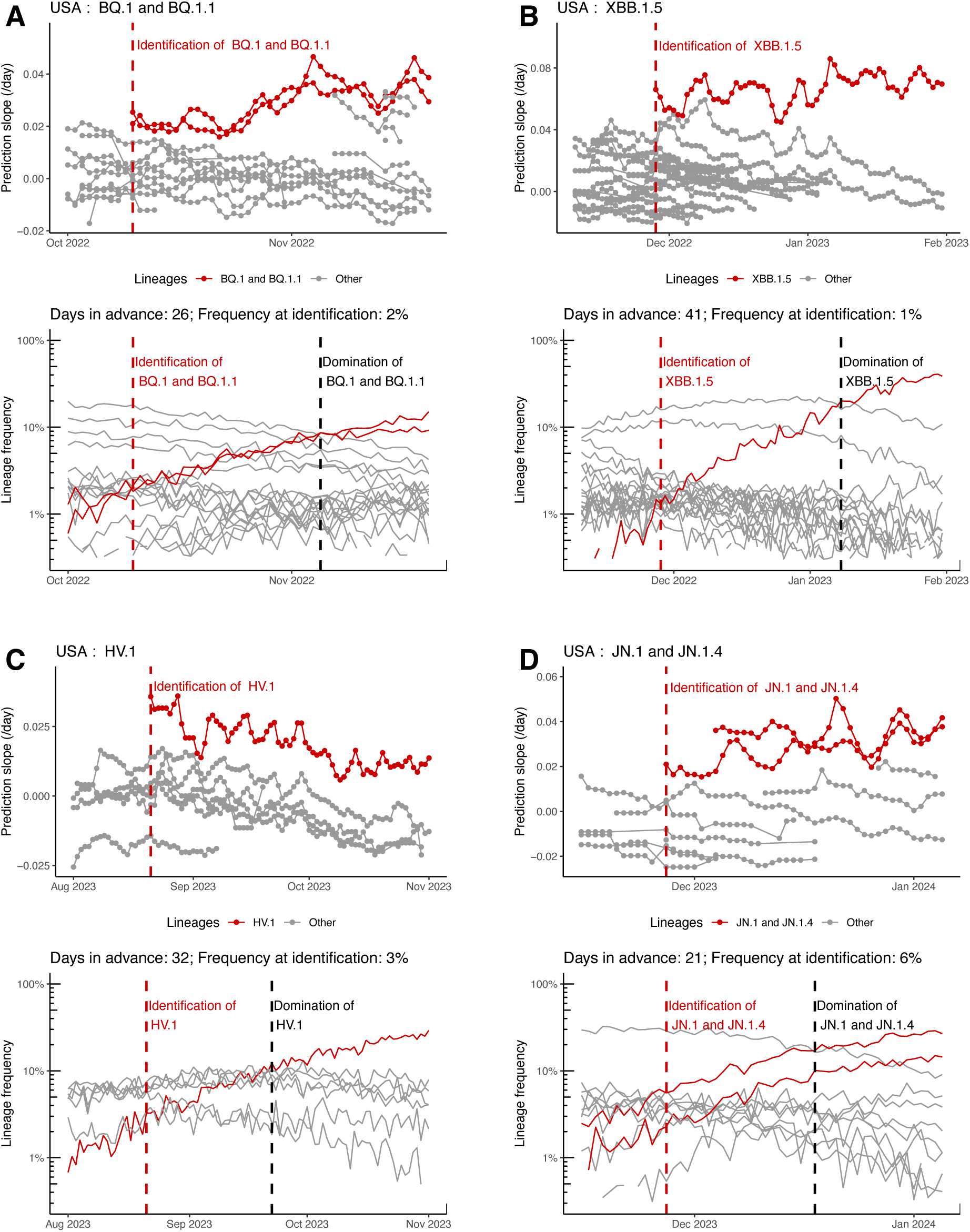
Identifying the rise of lineages in the US before they predominate. During the emergence of (A) BQ.1 and BQ.1.1, (B) XBB.1.5, (C) HV.1, and (D) JN.1 and JN.1.4, our model predicts the growth rates for all extant lineages. Focal and non-dominating lineages are colored in red and gray, respectively. In each panel, upper plots show the model-predicted slopes over time, and lower plots show the frequencies of each lineage. Red dashed lines show the day when our model identifies the future dominating lineage(s) as the most rapidly expanding, and black dashed lines show the time when the lineage(s) became dominant in the population. The number of days that identification precedes dominance of the lineage, and the frequency at the time of identification are indicated in the title of the lower panels.

**Figure 4:**
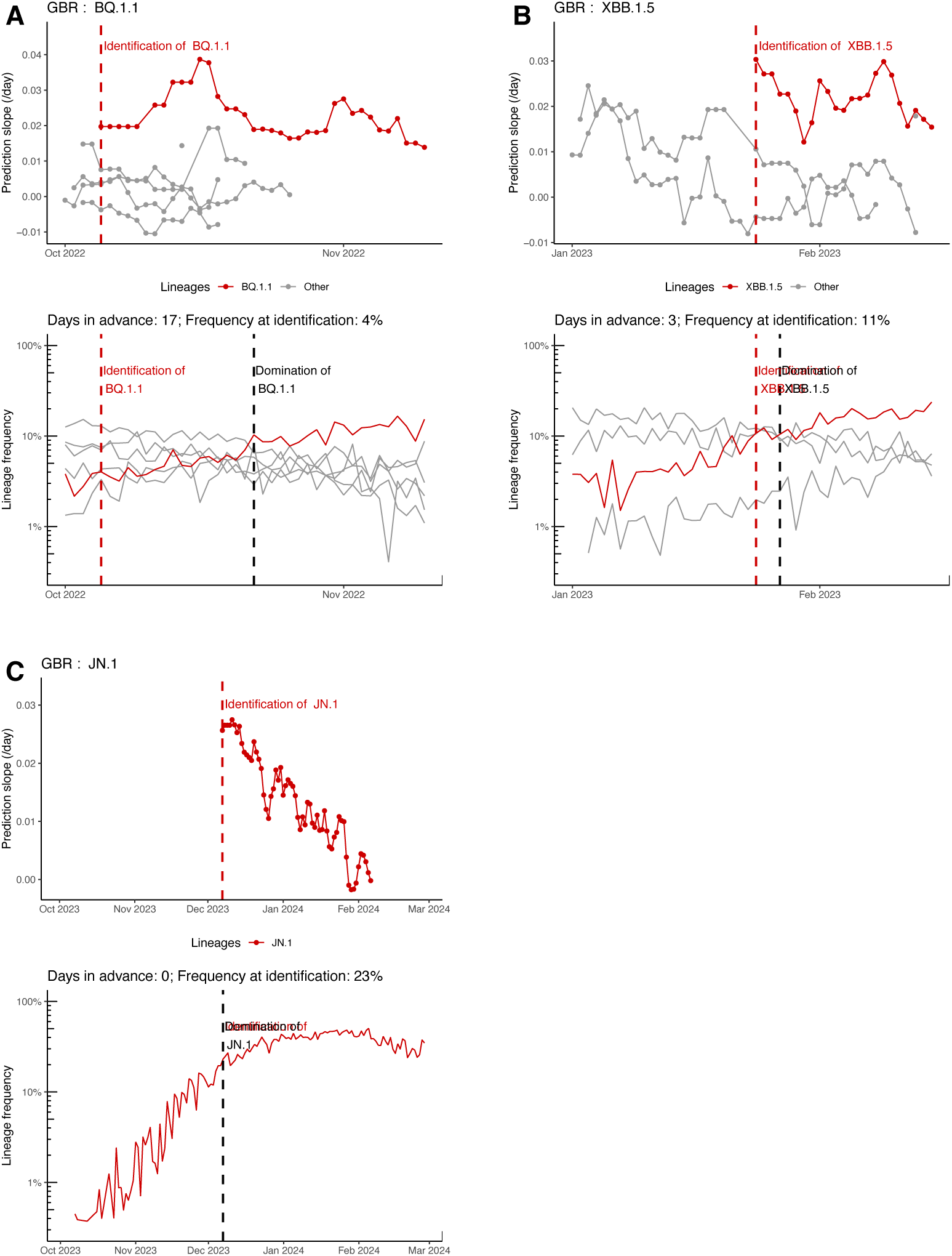
Identifying the rise of lineages in the UK before they predominate. Figure components are as in Fig. 3, but data are from the UK instead of the US.

The results from the UK raise the question whether using smaller submatrices would lead to earlier identification in situations where relatively fewer sequences are available. As we show in Fig. S12, for most lineages in our dataset, 30 sequences become available about 1-2 weeks earlier than 50 sequences do. We therefore tested our results using submatrices of size 30 sequences, and, for additional comparison, size 75 sequences. We found that smaller submatrix size leads to a somewhat weaker correlation within the training period and less accurate predictions for data from the testing periods (Fig. S11). We then applied the regression model derived from the smaller submatrices to retrospective real-time analysis. Indeed, identifications were made earlier using submatrices with 30 sequences, rather than 50, for most future-dominating lineages (6 out of 7) in the US and the UK (Figs. S5 and S6). In particular, the identification BQ.1 and HV.1 in the US and XBB.1.5 in the UK were made 7, 6 and 20 days earlier, respectively. This suggests the benefit of using smaller submatrices such as 30 ⇥ 30 for the purpose of identifying most rapidly expanding lineages, although their predictions of the slope are noisier.

Overall, these results show that our model is able to correctly identify rapidly expanding newly-emerged lineages. When the amount of sequencing is relatively large (e.g., the levels in the US throughout the period of analysis or in the UK between 2021 and 2022), identifications were made when the future dominanting lineages were still at a low frequency (<10%) and 3-6 weeks before they became dominant.

## Discussion

Early outbreak investigation involves estimating the growth rate, *r*, of a new outbreak or a newly emerging variant [21, 22, 23]. An estimate of *r* in turn allows for estimating a key epidemiological parameter, the reproductive number *R*, if the distribution of the generation time is known [24, 25]. Therefore, the sooner the growth rate of an infectious disease outbreak can be quantified and thus *R* can be estimated, the better can public health actions and personal decisions be made before the disease becomes widespread. In this work, we developed a graph theory based approach to process and analyze a large amount of pathogen genetic sequence data, and derived a model using two measures of subsets of genomic data—genetic diversity and connectivity—to estimate *r* for newly emerged variants. We showed how this approach can be used to effectively identify which of many existing variants are most likely to dominate in the future. Thus, our model could serve as an effective tool for pandemic monitoring purposes.

One major advantage of our approach is that it does not rely on lineage assignment. Although in our analysis we analyzed matrices within each lineage for convenience, in general sets of most-similar sequences can easily be identified without lineage name assignments. For example, when deploying our approach for pandemic monitoring purposes, one could first screen the new viral sequence data becoming available (on the order of thousands to tens of thousands per day) by first selecting the 50 most closely-related sequences for each sequence. Our PCA projection and regression model can then predict the rate of growth/decline of the population represented by the sequences. This will lead to identification of sequence clusters (among all other sequences within a parent lineage) that show the signature of rapid exponential growth, allowing early identification of the location of the outbreak and notification of evolving pandemic for public health officials.

Our approach does not rely on any specific epidemiological or evolutionary model, but it does have a strong intuitive basis. Closely related genetic sequences likely come from cases closely linked by transmission or an local outbreak, and the graph Laplacian of their pairwise genetic distance matrix thus reflects the rate of spread in a local population. We took measures derived from multiple submatrices in a country, however, to predict the growth rate. Thus, the result represents the average growth rate of many local outbreaks across the country. Compared to recent work that used sequence-based summary statistics (number of segregating sites or number of identical sequences) to predict the reproductive number [13, 15], we hypothesize that our approach has more power because the regression model is trained on the large amount of SARS-CoV-2 genomic data, and we include transmission network structure in the form of PC1 from the normalized Laplacian as additional information to make predictions. Compared to other work that estimates relative variant fitnesses [7, 26, 8], which can then be used to forecast variant proportions, out approach does not rely on lineage assignment (see above) nor a specific model of selection, and it can make predictions based on less data, sooner in an outbreak.

Our work has several limitations. First, we caution that although our PC1 measure of matrix connectivity serves as a good indicator of the rate of initial lineage expansion and final lineage decline, it is not clear how to interpret its value in the period in between. Investigating how PC1 changes during the whole period of time for each lineage, we found that PC1 during the period when the frequency stayed around the peak, when the rate of change is close to zero, mostly remained similar to values calculated during the outbreak phase for a prolonged period of time. This is especially true for rapidly expanding lineages during the outbreak phase (e.g., BA.1.1 and BA.2.12.1 in the US as shown in Fig. S9CD). This is because, when a population expands very quickly, coalescence events mostly likely occur near the time when the population size was small, at the beginning of the lineage expansion. This remains true later on when the population size becomes large and stops expanding. Therefore, the connectivity structure of the graph Laplacian (which is partially determined by the coalescence events) remains the same for long periods of time, leading to consistently large PC1 values and thus high estimated slopes even when the population size starts to saturate.

Our model is trained on data from only two countries, i.e. the US and the UK. It is not clear how transferable the parameter values are to data from other countries where sampling efforts are considerably lower. In addition, our model requires at least 50 sequences per day for several days to make reliable predictions of the growth rate, and at least 30 to effectively identify most rapidly growing lineages. This becomes a stringent criterion for early identification when the level of the sampling effort becomes low (e.g., in 2024 in the UK; Fig. 4). One potential solution is to analyze sequences collected in a few consecutive days to form submatrices. Further work is warranted to investigate the transferability of the model and its robustness to modifications that would require less data. Finally, when we test how early our model can identify future-dominating lineages (e.g. see Fig. 3), we used data collected retrospectively. In real time for those periods we analyzed, less sequences were available in the database, which means that we would be able to estimate the growth rate of newly emerged lineages from graph features at a later time than we showed in the figures, and at the same time, we may only identify a lineage becoming dominant in the population at a later time than we showed in these figures.

Overall, by selecting an appropriate unit of analysis and applying dimension reduction techniques that select for signatures of population growth, our approach can efficiently process large amount of data and at the same time make reliable predictions (of epidemic growth rate *r*) using a small amount of genomic data in a computationally efficient manner. More broadly, beyond estimating the early epidemic growth rate, we hypothesize that other applications of the graph Laplacian to pairwise genetic distance matrices could reveal additional features of the network structure of disease transmission. For example, future work could focus on extracting features of epidemic substructure and superspreading effects—two important epidemiological parameters—from submatrices, or on analyzing the utility of additional features from randomly-sampled sets of sequences (rather than sets of most-closely-related sequences, as we used). In principle, our approach should be equally applicable to other rapidly-evolving pathogens. In practice, one would need to train a similar PCA and regression model for a pathogen of interest, and potential for a given sampling regime. However, less data would be available for a newly-emerging pathogen, which could affect the stability of the PCA and/or regression components. Alternatively, one can determine whether the SARS-CoV-2 model and parameters we trained are transferrable across pathogens, or train a model using data simulated under an evolutionary-epidemiological process that reflects whatever information is known about the pathogen.

## Methods

### Genetic sequence data preparation

All available SARS-CoV-2 metadata and genomic sequences were downloaded from GISAID [19] on 2024-10-07. To avoid artifacts in recent data caused by a delay between the dates of sample collection and data upload, we used only sequences collected before 2024-07-01. All discussion of sequence dates below uses the collection date (when the person was sampled), not the submission date (when the sequence was provided to GISAID). We included data from the United States (country code USA) and the United Kingdom (country code GBR). Nextclade [20, version 3.8.2] was used to align each sequence to the Wuhan-Hu-1/2019 (MN908947) reference. Sequences with an invalid collection date or a Nextclade overall quality score of ‘bad’ were then excluded. We also ignored the first and last 100 positions in the genome, which are often N, and further excluded sequences that did not have at least 90% ACGT content at the remaining positions. The list of all sequences used in our analyses is provided at https://doi.org/10.55876/gis8.250128fy. Code for our analyses is provided at https://github.com/ruianke/CoV2-GenomicGraphTheory-PCA.

Our approach is founded on the expectation that a small collection of closely-related sequences will carry the signature of a recent outbreak. We therefore formed pairwise distance matrices for small subsets of sequences (‘submatrices’). To do this, we partitioned the data into sets for each day, for each Pango lineage [10] as reported by GISAID, for each country. For each day’s set, we formed 20 replicate submatrices by first choosing one sequence at random, and then including the *M* - 1 sequences with the smallest pairwise Hamming distance from it (`ape` function `dist.dna(…, method=‘raw’, pairwise.deletion=T)`; [27, 28]).

We repeated this for *M* = 30, 50, and 75, including only days with at least *M* sequences. Our main results use *M* = 50, which provides relatively early prediction and good accuracy, discussed further in the text.

### Graph Laplacian PCA to distinguish outbreak from non-outbreak

We hypothesized that the submatrices taken during the exponential lineage growth phase differ substantially from the submatrices from the decline phase. Our reasoning is that the structure of the submatrices should reflect past population dynamics of the virus population, analogously to how the structure of a phylogenetic tree reflects past population dynamics [17]. We therefore converted each submatrix to a Laplacian matrix, representing each sequence as a node in a graph and the pairwise distance between two sequences as the level of connection between two nodes. For each submatrix *W* , its graph Laplacian matrix *L* is *L* = *D* -*W* , where *D* is the degree matrix of *W* . The normalized distance Laplacian matrix *L* is obtained by dividing each entry of *L* by the vertex degree [29].

In graph theory, the eigenspectrum (the ordered list of eigenvalues) reflects the connectivity structure of the whole graph. Thus, the eigenspectrum of the Laplacian reflects the structure in the patterns in the pairwise distances. Previous work has shown that distinct transmission dynamics leave distinct patterns in phylogenetic trees [17] and that tree properties are visible in the Laplacian spectrum [30]. We therefore hypothesized that distinct transmission dynamics—in our case, the initial outbreak versus final decline of a variant—would be distinguishable from the Laplacian eigenspectrum. We obtained the eigenvalues of *L* and *L* using the `R` function `eigen()` [27]. Note that the eigenvalues are invariant with respect to the ordering of sequences in the pairwise distance submatrix, in contrast to approaches that treat the submatrix as an image [14]. We expected that these eigenvalues would exhibit distinct differences between small groups of sequences that did or did not come from a rapidly-growing outbreak.

To train the graph Laplacian component of our model, we took submatrices from variants that met specific criteria designed to identify those that had a clear outbreak phase (Fig. 1A). We considered the time period beginning with the rise of B.1.1.7 (2020-11-05 in GBR, 2021-01-05 in USA) and ending with the fall of BA.2 (2022-06-28 in GBR, 2022-07-11 in USA). This includes variants with a wide range of growth rates, from Alpha being relatively slow to Omicron being quite fast, while still allowing validation on later variants that were not used in training.

For all Pango variants, we identified the earliest and latest three days with enough data to form submatrices, for each country. We retained only those variants for which those are six unique days, all of which fall within the training time period. We additionally required that a variant have attained a frequency of at least 10% on at least one day in this time period. Our main results thus included 18 Pango-country combinations for training (Fig. 1B).

For each pairwise distance submatrix (each of six days, each variant, each of two countries; replicated 20 times), we formed the normalized graph Laplacian matrix and computed its eigenvalues (Fig. 1C). To reduce the dimensionality of the eigenspectrum, we then put that full collection of eigenvalues into a principal components analysis. Our subsequent analyses used the definitions of PC1 and PC2 from this single training PCA. Note that the PCA is unsupervised with respect to whether a submatrix comes from the rise (‘outbreak’) or fall (‘non-outbreak’) phase of the variant’s trajectory.

### Piecewise Linear Regression (PWLR) on frequency time series

We used piecewise linear regression (PWLR) to derive the best-estimate growth rate for each Pango lineage, to be used as ‘true’ values against which our submatrix-derived growth rates are compared. The data used included all sequence data available at the time of download; note that this means the analysis includes retrospective data, in order to obtain the best possible estimates of daily variant frequency. We assumed that variant frequency changes exponentially and thus fit a PWLR model to the log10-transformed variant frequency timeseries (Fig. 2A) using the ‘selgmented’ function in the ‘segmented’ package in R [31, 32, 27]. The ‘selgmented’ function first selects the number of breakpoints in the time series to break the time series into segments according to the BIC criterion and then performs linear regression on each of the segments.

### Regression model to explain growth rate

We used multivariate linear regressions to test combinations of various features extracted from submatrices to explain the best-estimate epidemic growth rate of the variant. We considered all Pango variants with sufficient data to form submatrices from the first 3 days of outbreak period and the last 3 days of the non-outbreak period, before October 2022; this is the same requirement as for the PCA, but over a longer time period so it includes many more lineages. For each submatrix, we computed its PC1 value using the PCA map determined from only early variants, described above, and the log10 of its nucleotide diversity (using the function `nuc.div()` in the R package pegas [27, 33]). We averaged the values of each of the features across all submatrices for each lineage to reduce noise (Fig. 2A). These mean values were used as predictor variables in the regression.

The growth rates for each lineage were estimated from the PWLR model as described above. Because patterns in the submatrices reflect recent population changes, we used the rate estimated for the date 7 days prior to each of the outbreak and non-outbreak sampling periods as the best-estimate growth rate for the period. Choice of the number of of days (within reasonable range) prior to the sampling period did not impact the final regression strongly. The resulting regression is:

*r* = -0.1772 + 0.0028 ⇥ *PC*1 - 0.0527 ⇥ log_10_(*nuc.div.*),

where *r* is the lineage growth/decline rate.

### Robustness to submatrix size

Our main analyses use submatrices of size *M* = 50 sequences. We also repeated the analyses using smaller and larger submatrices, *M* = 30 and 75. We found similar results for all submatrix sizes, with the regression model using PC1 and the logarithm of nucleotide diversity explaining a large fraction of variance in the true rates of change in viral frequency (compare Fig. S11 with Fig. 2).

Using smaller submatrices potentially allows for making predictions earlier when less data is available, but we found the performance was less accurate for *M* = 30 (*R*^2^ of the regression analysis is only 0.38), while remaining similar between *M* = 50 and 75 (*R*^2^=0.56 for both M values) (Fig. S11). We therefore considered *M* = 50 to be a good choice considering both accuracy and timely estimation.

## Data Availability

All data produced will be made available online at the time of publication.

## Acknowledgments

Research presented in this article was supported by the Laboratory Directed Research and Development program of Los Alamos National Laboratory under project numbers 20230830ER and 20240066DR.

This research used resources provided by the Darwin testbed at Los Alamos National Laboratory (LANL) which is funded by the Computational Systems and Software Environments subprogram of LANL’s Advanced Simulation and Computing program (NNSA/DOE).

We gratefully acknowledge all data contributors, i.e., the Authors and their Originating laboratories responsible for obtaining the specimens, and their Submitting laboratories for generating the genetic sequence and metadata and sharing via the GISAID Initiative, on which this research is based.

