## Supplementary Figures for "Estimating rates of SARS-CoV-2 lineage spread from graph theory analysis and data mining of genetic sequence data streams"

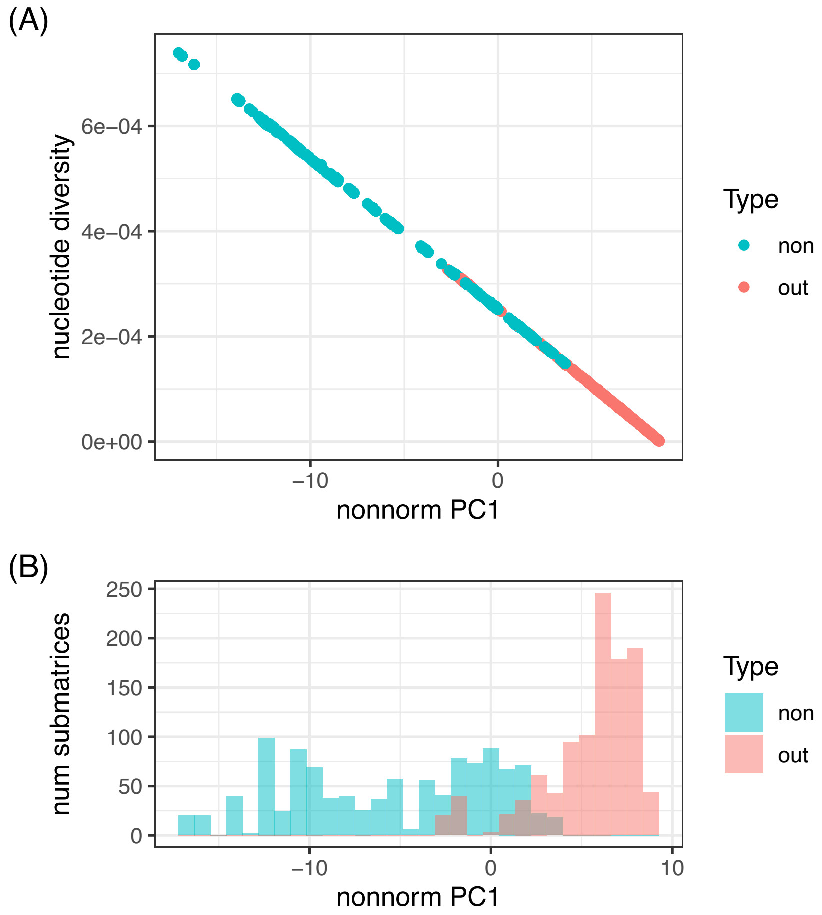


Figure S1: First principal component from the non-normalized Laplacian submatrices.

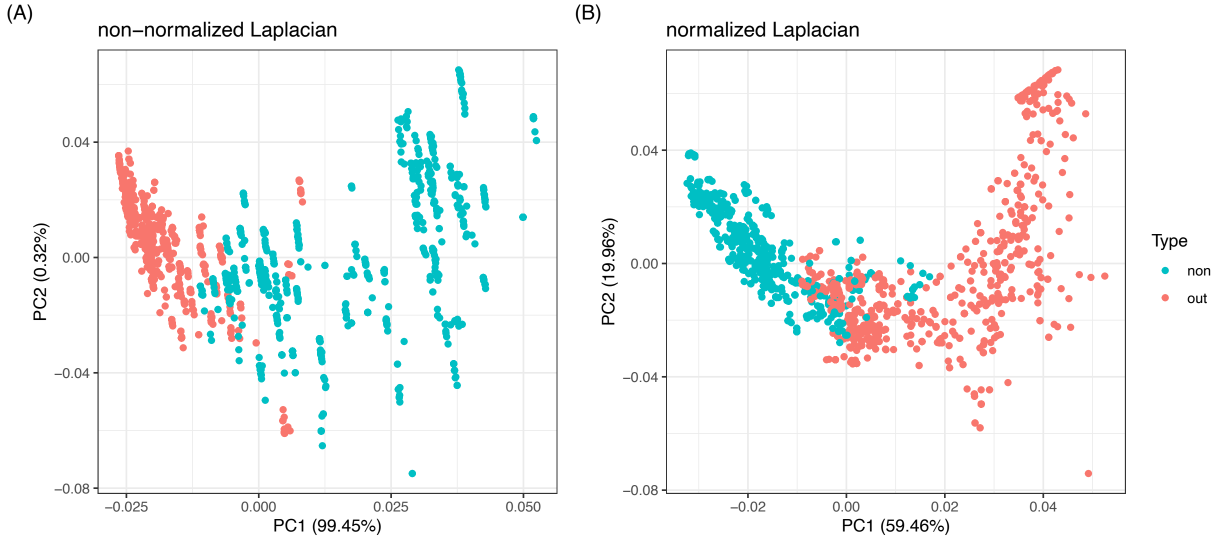


Figure S2: First and second principal components from the non-normalized and normalized Laplacian submatrices.

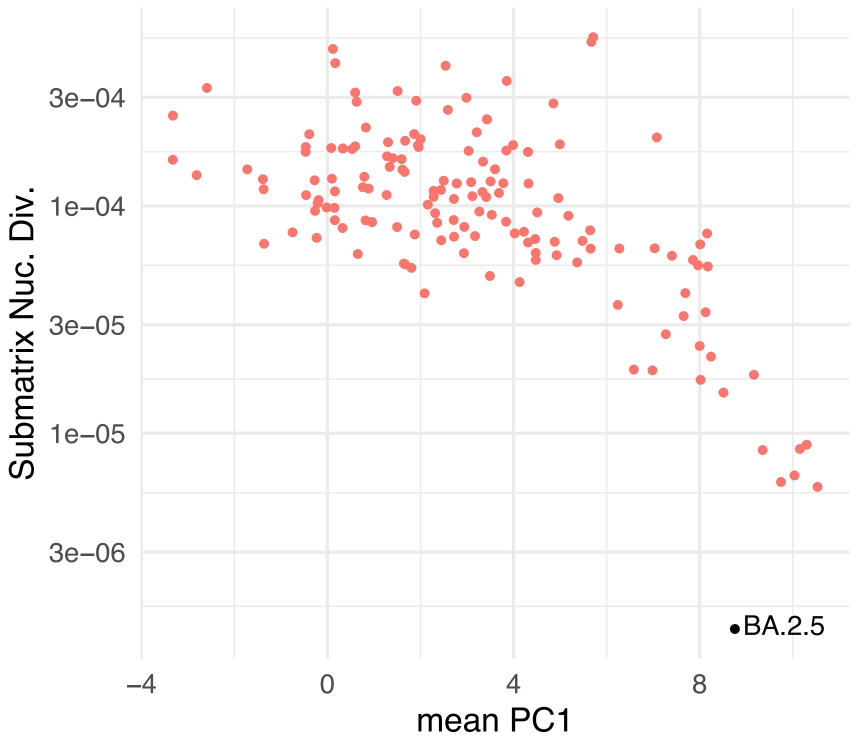


Figure S3: BA.2.5 has extremely low genetic diversity ($< 3 \times 10^{-6}$).

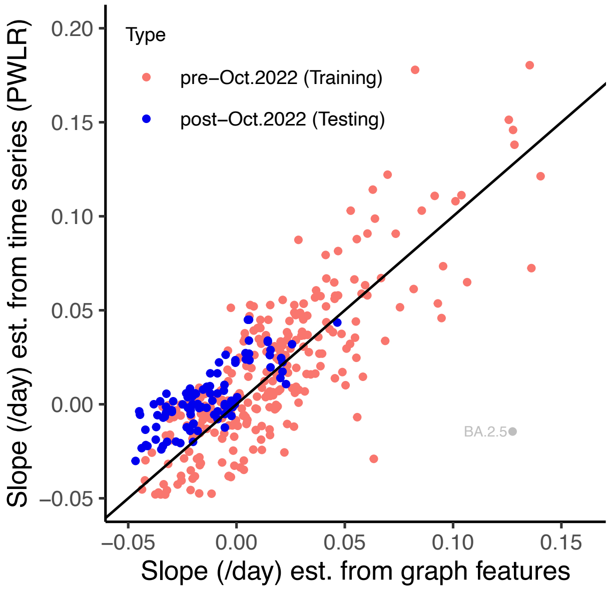


Figure S4: Linear regression model to predict epidemic growth rate using data from both the outbreak and the non-outbreak phases of each lineage. The slopes predicted from this model (horizontal axis) correlated well the slopes from the frequency timeseries (vertical axis). As in Fig. 2, pink dots represent the training data used to infer parameter values in the regression analysis, and blue dots represent the testing data used to test the regression predictions. BA.2.5 from the UK (gray dot) was excluded from the regression because of extremely low genetic diversity.

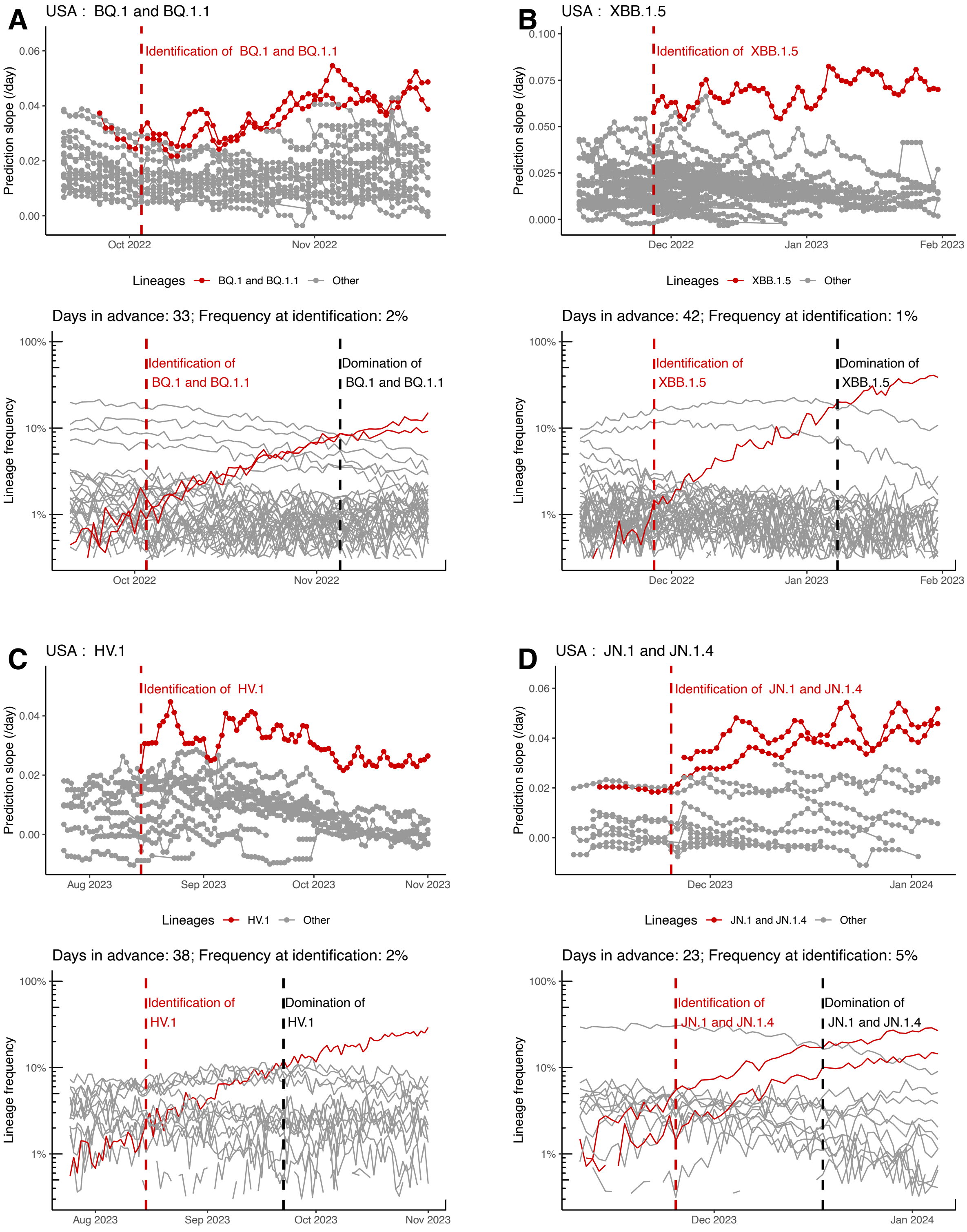


Figure S5: Model predictions of the rise of BQ.1, XBB.1.5, HV.1 and JN.1 lineages in the US using $30 \times 30$ matrices. The figure is plotted in the same style as Fig. 3 except using 30 sequences per submatrix.

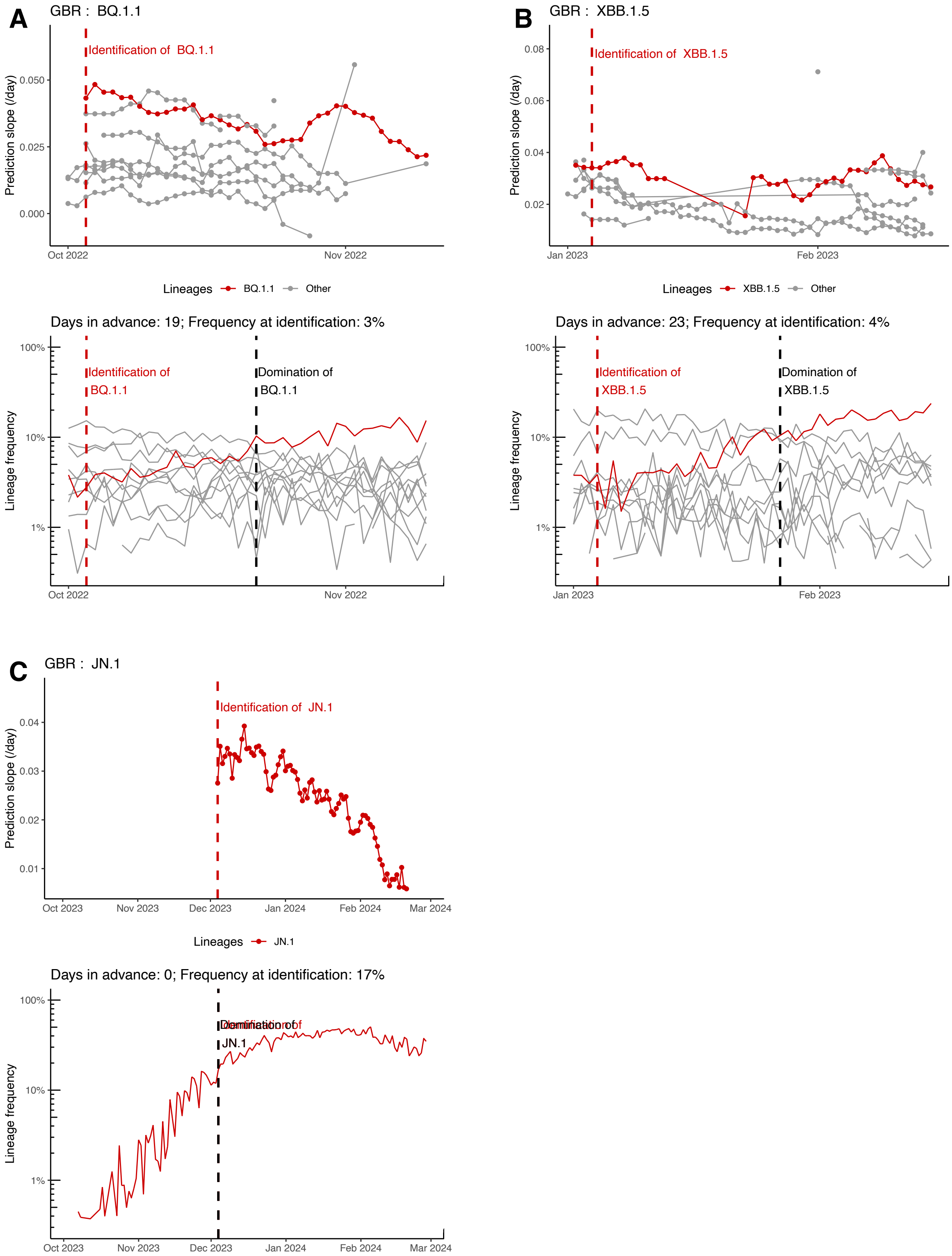


Figure S6: Model predictions of the rise of BQ.1, XBB.1.5 and JN.1 lineages in the UK using $30 \times 30$ matrices. The figure is plotted in the same style as Fig. 4 except using 30 sequences per submatrix.

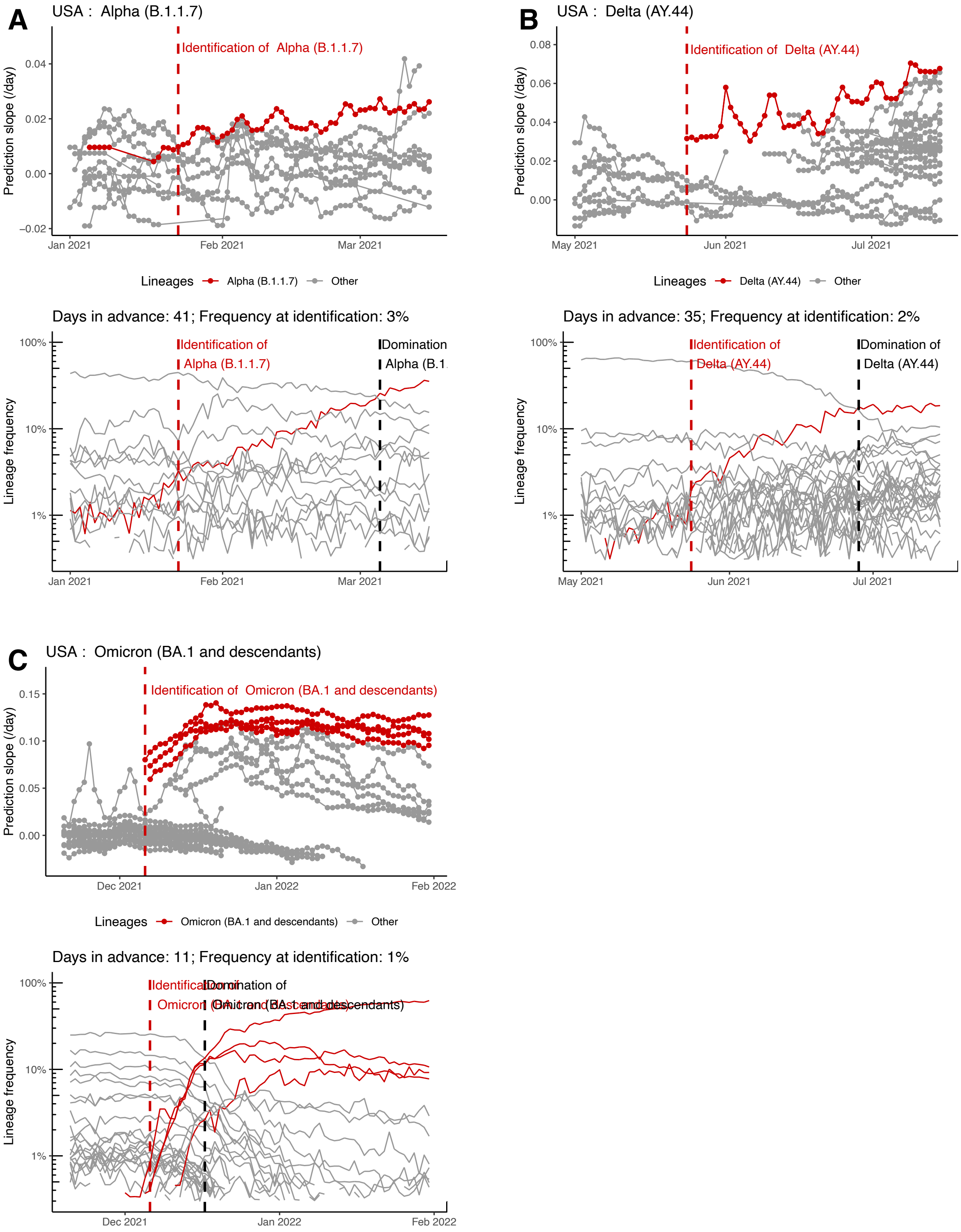


Figure S7: Model predictions of the rise of Alpha, Delta, Omicron (BA.1) in the US. The figure is plotted in the same style as Fig. 3 except using the training dataset.

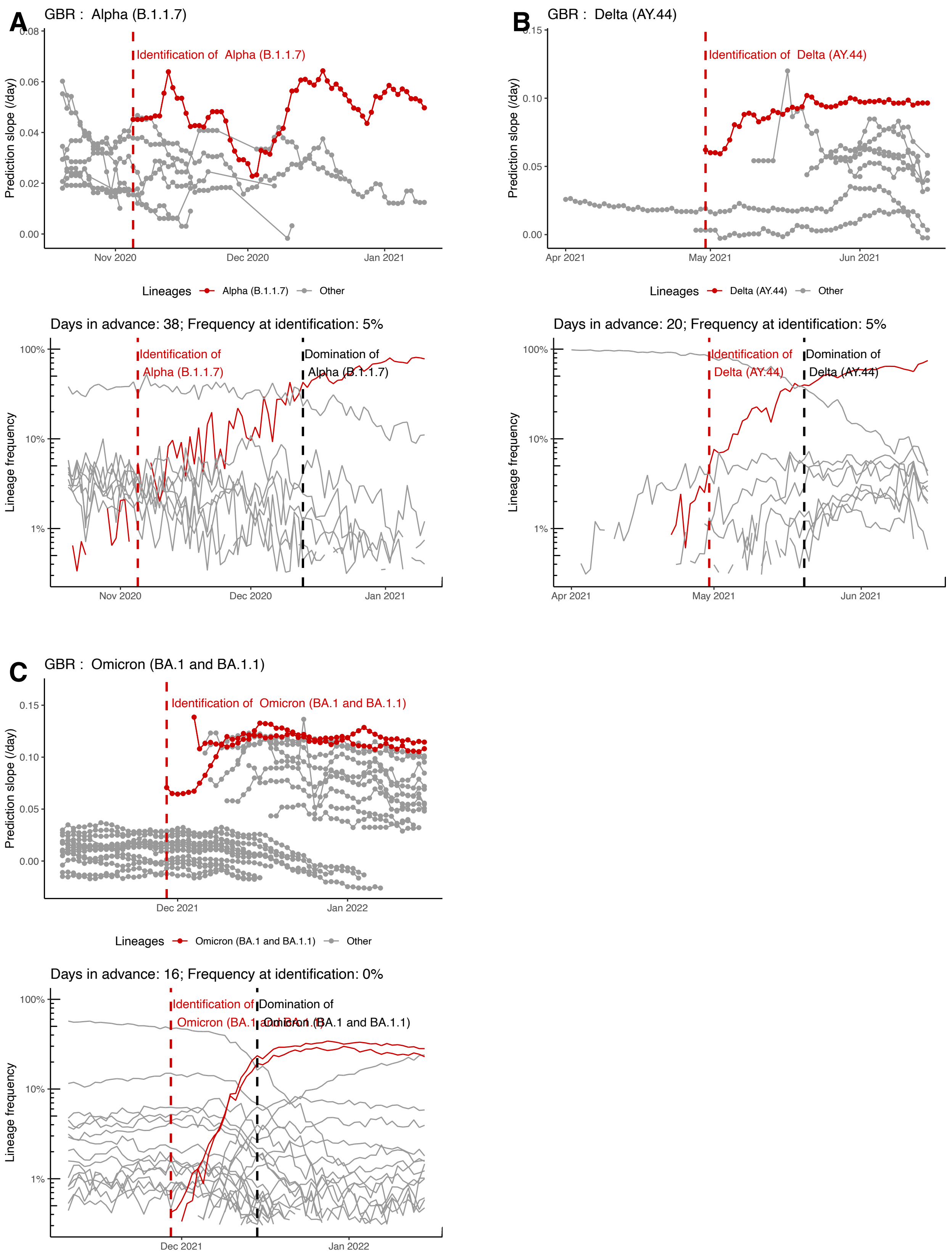


Figure S8: Model predictions of the rise of Alpha, Delta, Omicron (BA.1) in the UK. The figure is plotted in the same style as Fig. 4 except using the training dataset.

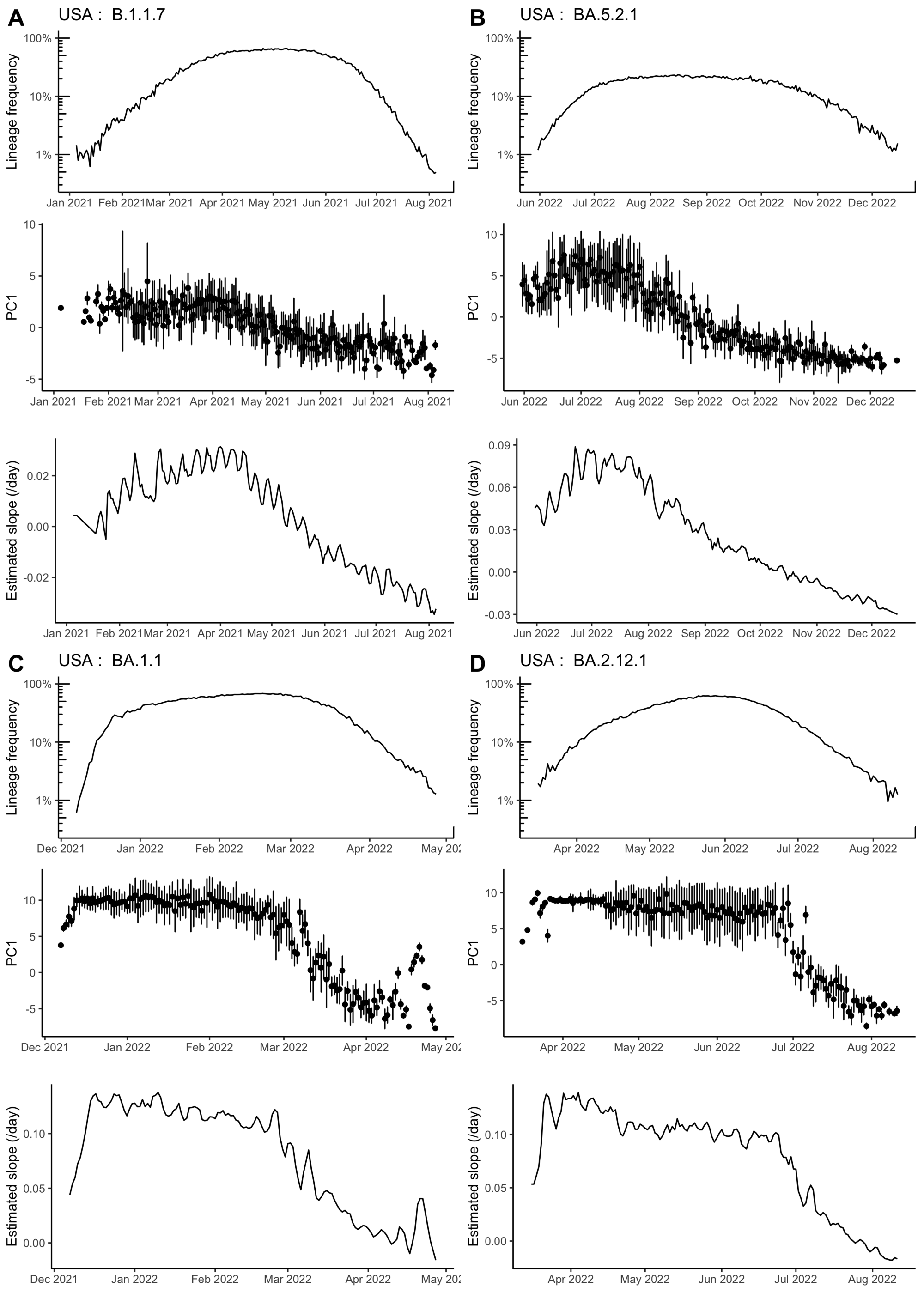


Figure S9: Lineage frequency, PC1 graph feature, and model-estimated growth rates for the full duration of four example lineages. Slopes estimated by our regression model are most accurate during the exponential growth and decline phases of the lineage (as also demonstrated in Fig. S4). During the periods when the lineages stayed dominate in the population, true growth rates are low but the estimated slopes are still high, possibility because of the large population size which led to no or minimal structural changes in the submatrices collected over this period.

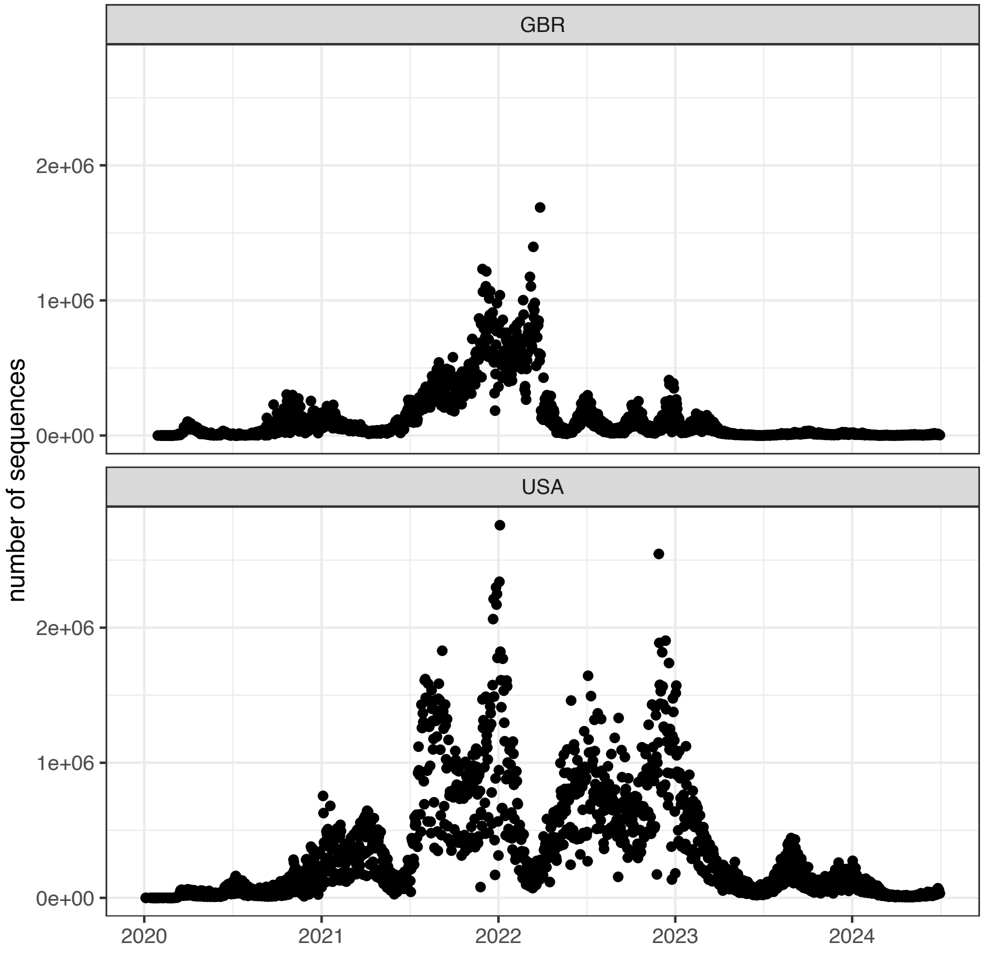


Figure S10: Total number of sequences over time, across all Pango in the UK (upper panel) and the US (lower panel).

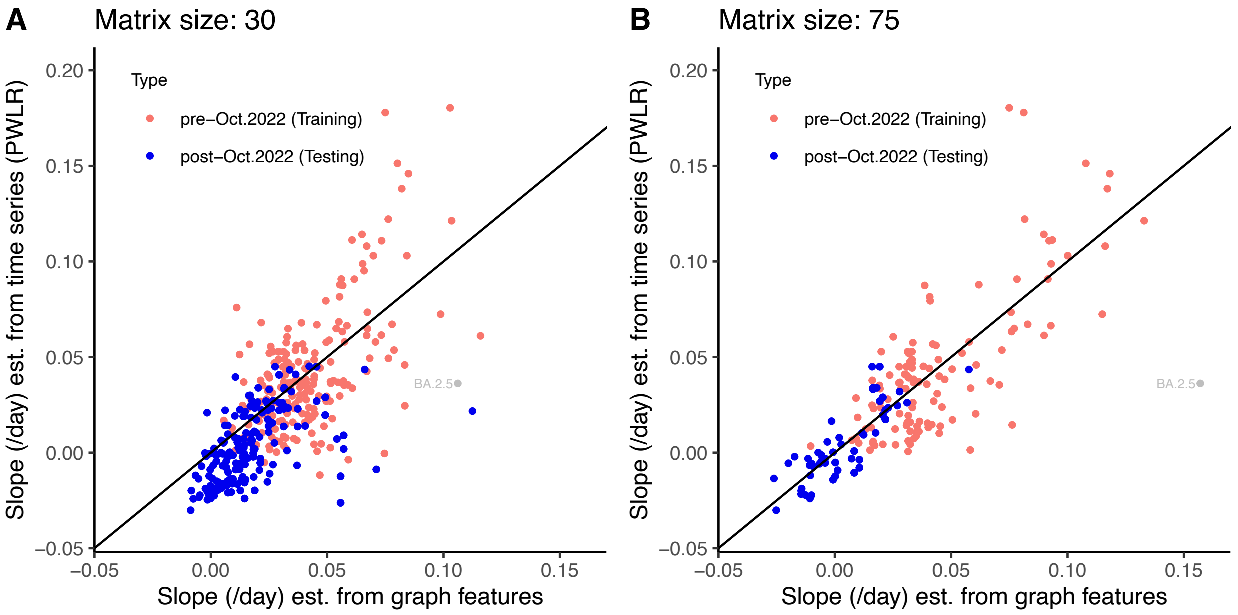


Figure S11: Linear regression model to predict epidemic growth rate using (A) smaller and (B) larger sub-matrices than our main results. In general, the slopes predicted from this model (horizontal axis) correlated well with the slopes from the frequency timeseries (vertical axis). The $R^2$ values are 0.38 and 0.56, for the $30 \times 30$ and $75 \times 75$ matrices, respectively. Note that the total number of data points used for regression and for testing differ between the two matrix sizes, because fewer lineages are available when using large matrix sizes. As in Fig. 2B, pink dots represent the training data used to infer parameter values in the regression analysis, and blue dots represent the testing data used to test the regression predictions. BA.2.5 from the UK (gray dot) was excluded from the regression because of extremely low genetic diversity.

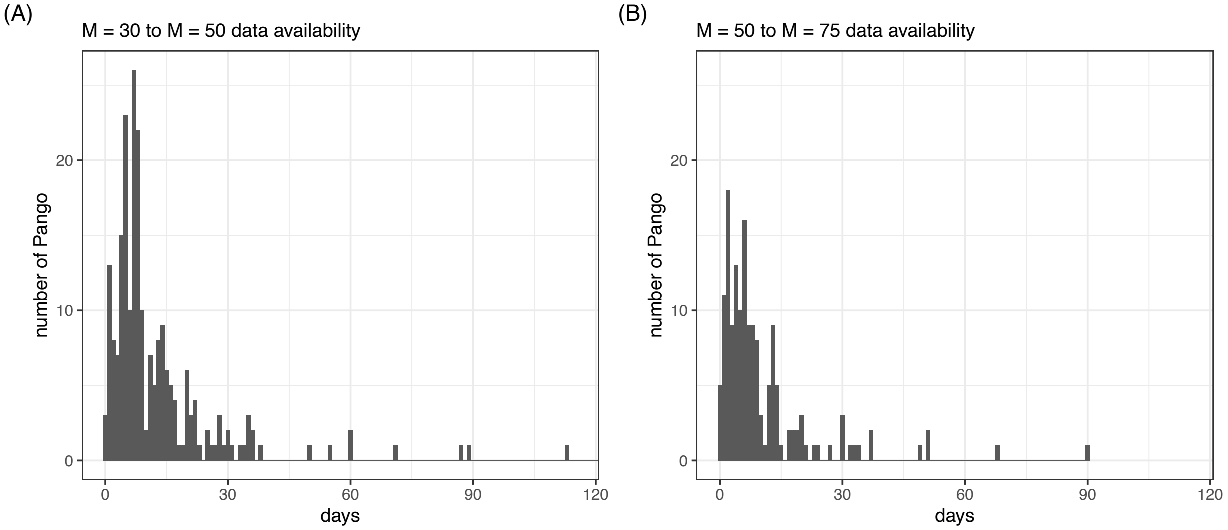


Figure S12: Prediction delay caused by requiring larger submatrices. For each Pango in each country, we identified the third day on which there was sufficient data to form a submatrix. The difference in these dates for two submatrix sizes, $M$, is shown as a histogram. Omitted from (A) is one value of 607 days. Additionally, 131 lineages were lost by requiring $M = 50$ rather than 30, and 200 lineages were lost by requiring $M = 75$ rather than 50.
